# Vaccine Effectiveness against Referral to Hospital and Severe Lung Injury Associated with COVID-19: A Case-Control Study in St. Petersburg, Russia

**DOI:** 10.1101/2021.08.18.21262065

**Authors:** Anton Barchuk, Mikhail Cherkashin, Anna Bulina, Natalia Berezina, Tatyana Rakova, Darya Kuplevatskaya, Oksana Stanevich, Dmitriy Skougarevskiy, Artemiy Okhotin

## Abstract

**Background:** Results of a randomised trial showed the safety and efficacy of Gam-COVID-Vac against COVID-19. However, compared to other vaccines used across the globe, the real-world data on the effectiveness of Gam-COVID-Vac, especially against the disease caused by Delta variant of concern, was not available. We aimed to assess the effectiveness of vaccination mainly conducted with Gam-COVID-Vac in St. Petersburg, Russia.

**Methods:** We designed a case-control study to assess the vaccine effectiveness (VE) against lung injury and referral to hospital. Self-reported vaccination status was collected for individuals with confirmed SARS-CoV-2 infection who were referred for initial low-dose computed tomography triage in two outpatient centres in July 3 – August 9, 2021 in St. Petersburg, Russia. We used logistic regression models to estimate the adjusted (for age, sex, and triage centre) VE for complete (>14 days after the second dose) vaccination. We estimated the VE against referral for hospital admission, COVID-19-related lung injury assessed with LDCT, and decline in oxygen saturation.

**Results:** In the final analysis, 13,893 patients were included, 1,291 (9.3%) of patients met our criteria for complete vaccination status, and 495 (3.6%) were referred to hospital. In the primary analysis, the adjusted VE against referral to hospital was 81% (95% CI: 68–88) for complete vaccination. The VE against referral to hospital was more pronounced in women (84%, 95% CI: 66–92) compared to men (76%, 95% CI: 51–88). Vaccine protective effect increased with increasing lung injury categories, from 54% (95% CI: 48–60) against any sign of lung injury to 76% (95% CI: 59–86) against more than 50% lung involvement. A sharp increase was observed in the probability of hospital admission with age for non-vaccinated patients in relation to an almost flat relationship for the completely vaccinated group.

**Conclusions:** COVID-19 vaccination was effective against referral to hospital in patients with symptomatic SARS-CoV-2 infection in St. Petersburg, Russia. This protection is probably mediated through VE against lung injury associated with COVID-19.

## Introduction

Vaccination proved to be an effective pharmaceutical control measure during the COVID-19 pandemic. More than 4 billion doses have been already administered, but real-world evidence is not equally available for all vaccines used globally. Even though the results from clinical trials of Gam-COVID-Vac [1, 2] suggested safety and efficacy similar to other COVID-19 vaccines used in population-based programmes across the globe [3, 4], this finding was met with scepticism and criticism [5–9]. Despite adoption of Gam-COVID-Vac in 49 countries, the independent safety data for this vaccine is available only from San Marino and Argentina [10, 11]. Further lacking is real-world evidence on Gam-COVID-Vac vaccine effectiveness (VE) from population-based studies.

A case-control study design and its modifications are the core methods to assess the effectiveness of vaccination in real-world settings [12]. Multiple case-control studies of COVID-19 vaccines have been published [13, 14]. The study that explored the VE of vaccines available in Scotland showed diminished protection against Delta VOC infection and hospital admission [15]. The COVID-19 pandemic in Russia has claimed more than 500,000 excess deaths by the spring of 2021 [16] while the lack of real-world evidence on the VE became a public health issue in Russia. Vaccination uptake in the country was undermined by vaccine hesitancy [17], partly driven by the lack of independent research exploring vaccine safety and effectiveness and failures to communicate the balance of benefits and harms of COVID-19 vaccination.

St. Petersburg is the second most populated city in Russia and the fourth in Europe. More than 45% of the population have contracted the SARS-CoV-2 infection by the end of April, 2021 [18]. However, only 10% received at least one dose of any vaccine by April, 2021 which is below the levels seen in other countries [18]. Low uptake by the spring of 2021 failed to stop the spread of the new Delta VOC in May–June 2021 in St. Petersburg. The spread of the Delta VOC, in turn, caused the rise in vaccination demand in the summer of 2021. As reported by the city government, by July 23, 2021, approximately 27% of the adult population of St. Petersburg received at least one dose of any vaccine, and 19% received two doses [19]. This increase in the vaccination uptake, consequently, has led to an increase in the absolute number of vaccinated individuals who reported to have a breakthrough infection or hospital admission. Such anecdotal evidence has caused further mistrust in the vaccination programme in absence of reliable scientific reports.

In this paper we present the first independent assessment of the VE in Russia. We designed a case-control study to assess the VE against referral to hospital and lung injury in the individuals who reported symptoms and were referred for initial computed tomography assessment in two outpatient centres in St. Petersburg, Russia.

## Methods

### Population and study design

St. Petersburg and other cities in Russia have established a triage service for symptomatic patients with COVID-19. Symptomatic patients with confirmed SARS-CoV-2 (using polymerase chain reaction (PCR) test) are referred to outpatient triage, including brief physical examination and low-dose computed tomography (LDCT). The decision about hospital admission is based on symptoms, i.e. shortness of breath, oxygen saturation, overall clinical condition, and lung injury assessed with LDCT. We designed a case-control study with “other patient” controls (test negative study) [20] to determine the VE against hospital referral and COVID-19 lung injury in symptomatic patients with confirmed SARS-CoV-2 infection, who were referred to the LDCT triage.

We retrospectively collected individual-level patient data from two outpatient triage centres of the Medical Institute named after Berezin Sergey (MIBS), a private medical facility contracted by the city government to provide triage service for nearly half of the city districts. All patients referred to LDCT triage underwent brief physical examination, including pulse oximetry. A local computed tomography score (CT-score) was implemented in Russia [21]. It has five gradations (0, 1, 2, 3, 4) which are related to the volume of involved lung segments (0, < 25%, 25–50%, 50–75%, 75–100%). It is used in the country to define the severity of COVID-19 lung injury and to triage patients. CT-score of 3 and 4 means more than 50% of lung volume involvement and is often used as an indication for the hospital admission. Patients with CT-score less than 3, i.e. less than 50% lung injury seen on the LDCT, and in the absence of severe symptoms, are normally sent to out-patient treatment or follow-up.

### Vaccination status and outcomes

In our primary analysis, cases were all patients referred to hospital after triage starting from July 3, 2021 till August 9, 2021. Controls were the patients referred to outpatient treatment and follow-up after the LDCT triage in the same period. The information about self-reported vaccination status was collected at the time of appointment to the LDCT. The patients reported the number of doses and the dates of vaccination, the type of vaccine data was not collected. The majority of vaccinated residents in St. Petersburg received Gam-COVID-Vac. The proportion of the population that received other vaccines was 10% or less in St. Petersburg [19, 22].

We used several definitions of the vaccination status. Complete vaccination status was assigned to the patients who had reported receiving two doses at least 14 days before the referral to the LDCT triage. Partial vaccination status was assigned to the patients who failed to meet the above criteria for complete vaccination, but had reported receiving one dose at least 14 days prior to the referral. In the secondary analyses, we used the CT-score [21] and oxygen saturation as the secondary outcomes to assess the VE against COVID-19 lung injury. Oxygen saturation was grouped in the ranges according to the NEWS2 score [23] which was well-validated in non-COVID-19 settings. It has four gradations (0, 1, 2, 3) which are related to the oxygen saturation in the following categories: >95, 94–95, 92–93, <92.

### Statistical analysis

We modeled our study plan following the WHO interim guidance to evaluate COVID-19 vaccine effectiveness [24]. We collected information on all patients referred to the LDCT triage, as most patients were sent for out-patient treatment and outpatient follow-up. We used unconditional logistic regression for our primary and secondary outcomes to estimate odds ratios (ORs) for vaccination status among cases and controls, which approximates ORs for the outcomes (hospital admission, different levels of lung injury, and decline in oxygen saturation) among the vaccinated and non-vaccinated patients. The VE was calculated as 100% × (1 *-OR*) adjusted for age (continuous variable), sex, and the triage LDCT centre. Our sample size of 495 hospitalized patients (cases), 13,398 non-hospitalized patients (controls), and 1,291 patients with the complete vaccination status (exposure level of 9.3%) provides 80% power to detect an odds ratio of 0.58 (or the VE of 42%) and 1.46 (or the VE of −46%) for hospitalization at the 5% alpha level in our unmatched case-control study. All standard errors and confidence intervals were adjusted for heteroskedasticity with the Huber-Eicker-White sandwich estimator. Finally, we investigated the relationship between the hospitalization and the thin plate regression spline of the patient age by vaccination status in a semiparametric logistic regression.

### Ethical considerations

The Ethics Committee of the MIBS approved the study on June 21, 2020. The Ethics Committee of the Pavlov First Saint Petersburg State Medical University approved the joint study of COVID-19 vaccine effectiveness in St. Petersburg on July 15, 2020. All research was performed following the relevant guidelines and regulations. All participants signed the informed consent upon referral to the LDCT triage. The joint study of COVID-19 vaccine effectiveness in St. Petersburg was registered at ClinicalTrials.gov (NCT04981405, date of registration — Aug 4, 2021). This publication covers the data collected at the two outpatient centres of the MIBS that contributed to the study data.

### Contribution and role of the funding source

The study team was entirely responsible for the study design and for collecting and analysing the data. There was no additional funding of the study provided.

## Results

Overall, 13,893 patients were included in the final analysis. Patients characteristics are presented in Table 1. Among all patients, 1,964 (14.1%) received at least one dose, 1,379 (9.9%) received two doses, 1,291 (9.3%) met our criteria for complete vaccination status, and additionally 448 (3.2%) met our criteria for partial vaccination status (one dose at least 14 days prior to the referral). 495 (3.6%) patients were referred to hospital after the LDCT triage (232 or 4.1% from the one outpatient triage centre and 263 or 3.2% from the other). The majority of referred patients (63.1%) had CT-score 3–4, or >50% lung involvement on LDCT. Patients referred to hospital were also older (66.1% were older than 60 years). Only 17 (3.4%) patients who were referred for hospital admission met the criteria for complete vaccination.

**Table 1.**
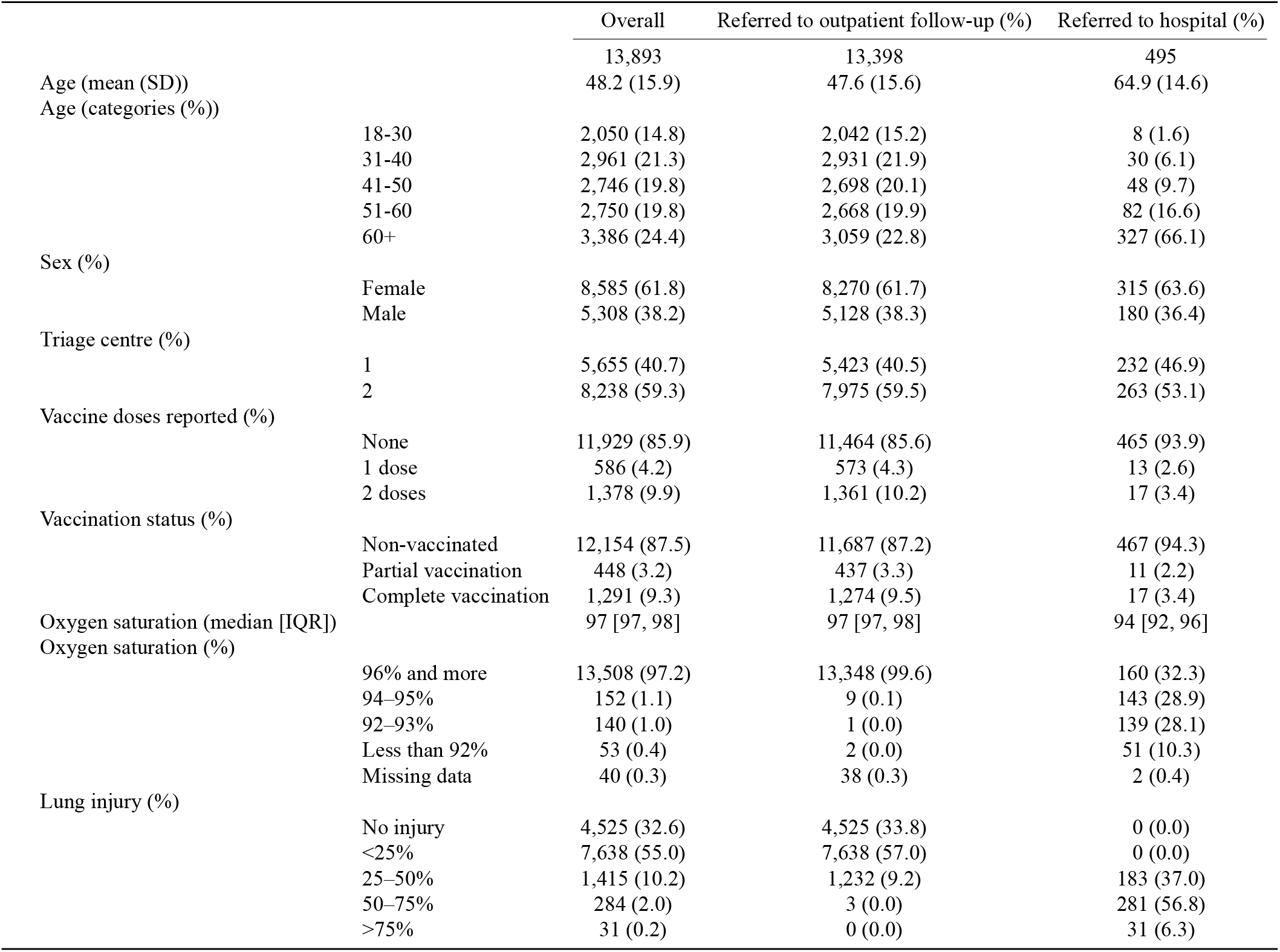
Characteristics of Persons with Covid-19 referred to the LDCT Triage.

In the primary analysis, the adjusted VE against referral to hospital admission was 81% (95% CI: 68–88) for complete vaccination (Table 2). The effect of the partial vaccination against referral to hospital was 35% (95% CI: −21–65).

**Table 2.** Effectiveness of Complete Vaccination Against Referral to Hospital, Lung Injury, and oxygen saturation decline.

|  |  | Crude OR (95% CI) | Crude VE (95% CI) | Adjusted OR (95% CI) | Adjusted VE (95% CI) |
| --- | --- | --- | --- | --- | --- |
| Triage result | Outpatient follow-up | 1.00 | — | 1.00 | — |
|  | Referral to hospital | 0.34 (0.21-0.55) | 66% (45-79) | 0.19 (0.12-0.32) | 81% (68-88) |
| Lung injury | No signs of lung injury | 1.00 | — | 1.00 | — |
|  | Any signs of lung injury | 0.64 (0.57-0.72) | 36% (28-43) | 0.46 (0.40-0.52) | 54% (48-60) |
|  | More than 25% | 0.60 (0.48-0.72) | 41% (28-52) | 0.36 (0.29-0.45) | 64% (55-71) |
|  | More than 50% | 0.42 (0.26-0.69) | 58% (31-74) | 0.24 (0.14-0.41) | 76% (59-86) |
| Oxygen saturation | 96% and more | 1.0 | — | 1.0 | — |
|  | Less than 96% | 0.47 (0.28-0.78) | 53% (22-72) | 0.30 (0.18-0.51) | 70% (49-82) |
|  | Less than 94% | 0.58 (0.32-1.07) | 42% (-7-68) | 0.39 (0.21-0.72) | 61% (28-79) |
|  | Less than 92% | 0.58 (0.18-1.85) | 42% (-85-82) | 0.34 (0.10-1.12) | 66% (-12-90) |

Crude and adjusted ORs and VE against lung injury following the LDCT assessment and decline in oxygen saturation is presented in Table 2. Only a few patients had more than 75% of lung involvement and none in the vaccination group, so we calculated the VE for the combined category, which included patients with more than 50% of lung involvement. The VE increased with the lung injury categories, from 54% (95% CI:48–60) against any signs of lung injury to 76% (95% CI:59–86) against more than 50% lung injury. The VE against different levels of oxygen saturation decline was consistent (Table 2).

The VE against hospital admission was more pronounced in women (84%, 95% CI: 66–92) compared to men (76%, 95% CI: 51–88) and older age groups (77%, 95% CI: 62–86) (Table 3). We observed a sharp increase in the probability of hospital admission with age for non-vaccinated patients in relation to an almost flat relationship between age and the probability of hospital admission for the completely vaccinated group of patients (Figure 1). There was no difference in the VE by the LDCT triage centre. Crude VE estimates were lower compared to the adjusted VE.

**Table 3.** Effectiveness of Complete Vaccination Against Referral to Hospital, According to Age Group, Sex, and Triage Centre.

|  |  | Crude OR (95% CI) | Crude VE (95% CI) | Adjusted OR (95% CI) | Adjusted VE (95% CI) |
| --- | --- | --- | --- | --- | --- |
| Sex | Female | 0.26 (0.13–0.53) | 74% (47–87) | 0.16 (0.08–0.34) | 84% (66–92) |
|  | Male | 0.46 (0.24–0.91) | 54% (9–76) | 0.24 (0.12–0.49) | 76% (51–88) |
| Age | 18–49 | 0.43 (0.10–1.74) | 57% (-74–90) | 0.37 (0.09–1.50) | 63% (-51–91) |
|  | 50 and older | 0.23 (0.13–0.38) | 77% (62–87) | 0.18 (0.11–0.31) | 82% (69–89) |
| Triage centre | 1 | 0.34 (0.16–0.72) | 66% (28–84) | 0.21 (0.10–0.46) | 79% (54–90) |
|  | 2 | 0.35 (0.18–0.65) | 65% (35–82) | 0.18 (0.09–0.36) | 82% (64–91) |

**Figure 1.**
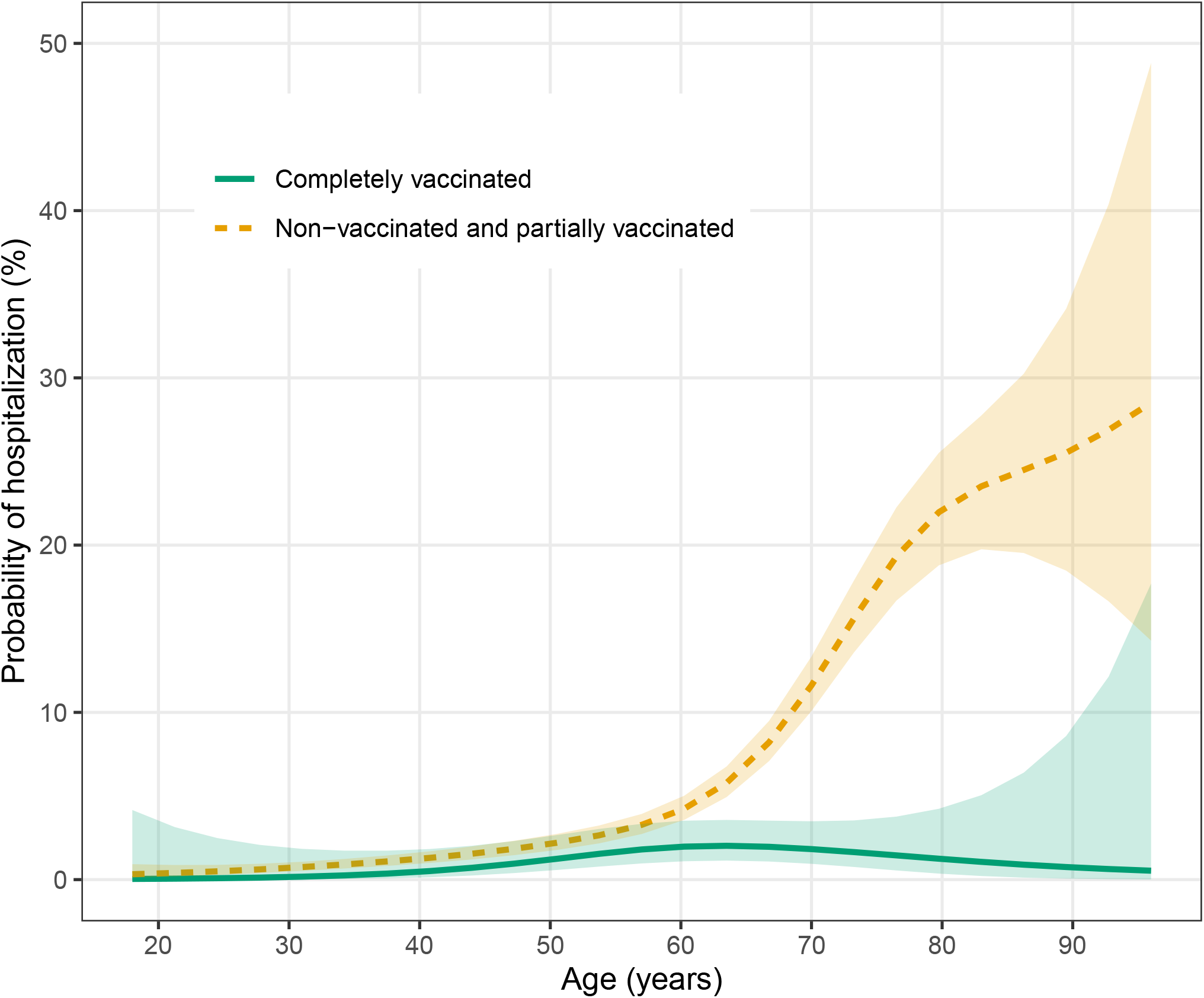
Probability of Referral to Hospital, According to Age and Vaccination Status (Shaded Areas Are 95% CI).

## Discussion

This is the first study examining the real-world effectiveness of COVID-19 vaccination in Russia, and one of the first studies globally to provide information about the vaccine protective effect against COVID-19-associated lung injury. Vaccination showed 81% (95% CI: 68–88) protection against referral to hospital in patients with symptomatic SARS-CoV-2 infection during the third wave of COVID-19 pandemic caused by Delta VOC in St. Petersburg. It is quite similar to the VE against Delta VOC reported for other COVID-19 vaccines [15]. Previously, only in-vitro data about reduced virus-neutralizing activity of Gam-COVID-Vac was published [25], but it was not clear how laboratory findings convert into clinical practice.

The clear strength of this study is the availability of independent lung injury assessment using the LDCT. The radiologists did not have information on the vaccination status, while the computed tomography was applied as a triage method for all symptomatic patients. The protective effect of vaccination was consistent against all grades of lung injury. We did not observe 75% or more lung tissue involvement during the LDCT in the group of fully vaccinated patients, suggesting strong and consistent protective effect. Protective effect was also observed against the decline in oxygen saturation. It was clear that both secondary outcomes (lung injury and decline in oxygen saturation) are closely related, but the oxygen saturation and LDCT were independently evaluated at the triage centres. Both factors are independently influencing the decision to refer to hospital, thus explaining the greater VE against hospital admission than for CT-score and oxygen saturation alone. These results further support the findings of the primary analysis.

Since all the patients in our study had COVID-19, our results provide specific information on vaccination impact on disease severity and its clinical course. When breakthrough infections are evolving as a cause of major concern, our data is assuring that the vaccine effect is going beyond the risk of contracting the infection. It shows that vaccination significantly diminishes disease severity and protects the lungs from virus-induced injury.

While SARS-CoV-2 is likely converting from a pandemic virus to an endemic one, the role of vaccination is also evolving from pandemic control to damage control. Our data shows that it is feasible. Infection fatality rate for COVID-19 is progressively increasing with age in St. Petersburg [18]. Our data shows that it is an effective way of protecting the most vulnerable elderly persons from devastating consequences of the pandemic.

Lung injury is not only observed in COVID-19 but also in many other respiratory viral infections [26]. Lung injury leading to pneumonia and acute respiratory distress syndrome is a major cause of morbidity and mortality in these diseases. Lung imaging with computed tomography were used for assessment and risk stratification in COVID-19 [27]. However, in the United States and Europe computed tomography was discouraged by the professional societies as the primary imaging modality in mild COVID-19 due to high cost and transmission risk with emphasis placed on clinical assessment instead [28]. In Russia, due to less developed primary care and the low cost of the LDCT, it became the primary imaging modality used for COVID-19 triage. This offered the unique opportunity to assess the VE against lung injury detected with LDCT.

Partial vaccination yields especially unreliable VE estimates due to low prevalence. The VE against Delta VOC for one dose of vaccine was lower than for two doses, and the effect did not clearly manifest until at least 28 days after the first one [15]. We obtained similar results in our study. The point estimate for partial vaccination was 34%, but the confidence interval for the corresponding OR included unity, suggesting inadequate sample size due to the minimum detectable VE of 70% for the partial vaccination exposure as our power calculations show. It should be noted that in Russia the recommended 21-day interval between the first and the second dose of Gam-COVID-Vac is usually strictly followed.

There are several important limitations of our study. The small number of vaccinated prevents us from conducting an adequately powered stratified analysis or estimating VE for one dose. It is also making the estimates less precise than ideal. We did not have the information on the vaccine type, so the estimated VE represent an average effect of vaccination in St. Petersburg. However, in its response to an inquiry by a member of St. Petersburg Legislative Assembl? city Health Committee revealed that, by July 23, 2021, in St. Petersburg, among 1,227,496 individuals who received at least one vaccine dose, 1,178,266 or 96% received Gam-COVID-Vac, and the rest is distributed between the other two vaccines (EpiVacCorona — 21,943 or 1.7% and KoviVac 27,287 or 2.2%) [19]. October 2021 survey showed that only about 10% of responders in St. Petersburg received a vaccine other than Gam-COVID-Vac [22]. It is safe to assume that our study approximates the effectiveness of Gam-COVID-Vac. The effectiveness for EpiVacCorona and KoviVac would be difficult to derive from observational case-control studies due to low uptake.

Another important limitation of our study is possible referral bias. Patients were referred to the triage centres if they had positive PCR tests and disease symptoms, but the decision to refer was left at the physician’s discretion. We cannot rule out that the vaccination status was influencing this decision. If this is the case, in our data we would observe more severely ill vaccinated individuals than the unvaccinated ones. Alternatively, the physicians may fully rely on vaccine protection and never refer the vaccinated patients to the triage. However, we observed a considerable number of vaccinated individuals at the triage centres. Therefore, we believe that the referral bias would result in underestimation of the VE. Vaccination could also influence patients’ medical care seeking behaviour. We assume that self-assurance on vaccine protection would predominantly prevent patients with mild disease from seeking medical care, and this would also lead to underestimation of VE against severe disease in our study.

The study results may be also biased if the triage referred symptomatic patients to hospital based on their vaccination status. To the best of our knowledge, this decision was solely based on physical examination and the LDCT results. The vaccination status was collected for research purposes, was not a part of any official medical records. There is little reason to believe that the vaccine influenced the decision to refer the patient to hospital. There was a slight difference between triage centres in the proportion of patients referred symptomatic patients to the hospital. Still, it was consistent with the difference in the proportion of patients with severe lung injury and unlikely to reflect inconsistency between the centres.

Finally, the data we used in our study was collected at the triage centres and not in hospitals. Even though all patients triaged for inpatient treatment were followed-up by the LDCT centre’s personnel from the moment they were referred to the hospital till hospital admission, we do not know the length or the outcome of the hospital admission itself. It is also possible that the patients triaged for outpatient treatment were later admitted. Our study covers only one point in their disease course and traces only the decisions made in the triage centres.

Additional observational studies are needed to assess the vaccination effectiveness against SARS-CoV-2 infection and COVID-19 associated death. Given the limitations of our data, we were only able to estimate the protective effect against lung injury and referral to hospital. However, it is safe to assume that those outcomes pose as a surrogate for COVID-19-associated death. Our study does not provide any information on the vaccine safety, but at least for Gam-COVID-Vac, the safety data are available from independent sources [10, 11].

In conclusion, we showed that COVID-19 vaccination is effective against referral to hospital in patients with symptomatic SARS-CoV-2 infection in St. Petersburg, Russia. The protection against hospital admission is probably mediated through protection against lung injury associated with COVID-19. Real-world evidence on the VE against COVID-19 should be integrated into population-based vaccination programmes to negotiate the balance of benefits and harms of this effective COVID-19 control measure, that is likely gaining even more importance in the light of gradual ceasing of non-pharmaceutical interventions against COVID-19 and calls for a “return to normality”.

## Data Availability

Study data and code is available online (https://github.com/eusporg/spb_covid_study20).

## Authors’ contributions

Anton Barchuk, Artemy Okhotin, Mikhail Cherkashin, and Oksana Stanevich conceived the study. Anton Barchuk, Artemy Okhotin, Mikhail Cherkashin, and Oksana Stanevich coordinated the study. Anton Barchuk, Artemy Okhotin, Dmitriy Skougarevskiy, and Anna Bulina drafted the first version of the manuscript. Mikhail Cherkashin, Natalia Berezina, Tatyana Rakova, Darya Kuplevatskaya, and Oksana Stanevich contributed to drafting sections of the manuscript. Mikhail Cherkashin, Natalia Berezina, Tatyana Rakova, Darya Kuplevatskaya collected data for analyses. Anna Bulina, Dmitriy Skougarevskiy, and Anton Barchuk did data analyses. All authors participated in the study design, helped to draft the manuscript, contributed to the interpretation of data, and read and approved the final manuscript.

## Acknowledgements

We thank Alexandra Vasilieva, Alla Samoletova, and Maria Batygina (European University at St. Petersburg) for the science communication and for administrative support. We also thank Evgeny Bakin for his insightful comments and Ivan Moiseev for study support. We thank the nurses and other personnel of the MIBS. We also thank all study participants.

